# Early changes in immune cell metabolism and function are a hallmark of sleeve gastrectomy: a prospective human study

**DOI:** 10.1101/2020.07.31.20161687

**Authors:** Tammy Lo, Eleanor J. M. Rudge, Robert P. Chase, Renuka Subramaniam, Keyvan Heshmati, Elizabeth M. Lucey, Alison M. Weigl, Otatade J. Iyoha-Bello, Chelsea O. Ituah, Emily J. Benjamin, Seth W. McNutt, Leena Sathe, Leanna Farnam, Benjamin A. Raby, Ali Tavakkoli, Damien C. Croteau-Chonka, Eric G. Sheu

**Author notes:** Co-supervised this work (Damien C. Croteau-Chonka, Eric G. Sheu).

## Abstract

**Objective:** To characterize longitudinal changes in blood biomarkers, leukocyte composition, and gene expression following laparoscopic sleeve gastrectomy (LSG).

**Background:** LSG is an effective treatment for obesity, leading to sustainable weight loss and improvements in obesity-related co-morbidities and inflammatory profiles. However, the effects of LSG on immune function and metabolism remain uncertain.

**Methods:** Prospective data was collected from 23 enrolled human subjects from a single institution. Parameters of weight, co-morbidities, and trends in blood biomarkers and leukocyte subsets were observed from pre-operative baseline to one year in three-month follow-up intervals. RNA-sequencing was performed on pairs of whole blood samples from the first six subjects of the study (baseline and three months post-surgery) to identify genome-wide gene expression changes associated with undergoing LSG.

**Results:** LSG led to a significant decrease in mean total body weight loss (18.1%) at three months and among diabetic subjects a reduction in HbA1c. Improvements in clinical inflammatory and hormonal biomarkers were demonstrated as early as three months after LSG. A reduction in neutrophil-lymphocyte ratio was observed, driven by a reduction in absolute neutrophil counts. Gene set enrichment analyses of differential whole blood gene expression demonstrated that after three months, LSG induced transcriptomic changes not only in inflammatory cytokine pathways but also in several key metabolic pathways related to energy metabolism.

**Conclusions:** LSG induces significant changes in the composition and metabolism of immune cells as early as three months post-operatively. Further evaluation is required of bariatric surgery’s effects on immunometabolism and consequences for host defense and metabolic disease.

## Introduction

Obesity is recognized as a chronic and systemic inflammatory disease^1^. A positive correlation between obesity and cellular immune dysregulation has been widely documented over the past decade, characterized by an increase in circulating levels of cytokines and interleukins^2–5^.

Since the original discovery by Hotamisligil *et al*.^2^ in 1993 demonstrating that cytokine levels are elevated in the adipose tissue of diabetic obese mice, and its neutralization improves glucose handling, evidence is accumulating that immune pathways regulate metabolic homeostasis. This chronic inflammation in obesity, due to oxidative stress in adipocytes, facilitates the macrophage infiltration cascade and atypical cytokine production leading to increased acute-phase proteins^4,5^. The activation of immune signaling pathways upsets multi-organ homeostasis especially in liver, brain and pancreas, and is responsible for the subsequent metabolic dysfunctions^3,6^.

With the incidence of obesity becoming pandemic^7^, there is a growing interest to investigate the relationship between bariatric surgery and its beneficial effects on immunometabolism. It is now widely accepted that laparoscopic sleeve gastrectomy (LSG) is a safe and effective stand-alone procedure^8,9^.

While it is rapidly increasing in popularity both as a metabolic and weight loss surgery of choice, LSG also improves systemic inflammatory profile^10^. A prospective study comprising of 22 participants with impaired glucose homeostasis undergoing LSG observed an improvement in leptin, C reactive protein (CRP), and interleukin-6 (IL-6) six months following surgery^11^.

To date, there is a lack of evidence demonstrating the impact and timing of LSG on immune cell metabolism and function. The goal of this prospective study was to characterize longitudinal changes in immunometabolic blood biomarkers over 12 months in human subjects with obesity prior to and following LSG. We also examined the relationship of surgical weight loss with genome-wide gene expression changes in peripheral blood leukocytes. We hypothesized that LSG restores normal immune function by inducing changes in immune cell metabolism and composition.

## Methods

### Study population

Subjects were identified and recruited prospectively from patients undergoing laparoscopic sleeve gastrectomy (LSG) between January 2017 and July 2019 at Brigham and Women’s Hospital (BWH) (Boston, MA). A multidisciplinary clinical team was involved and endorsed the indication for bariatric surgery. Decisions to undergo sleeve gastrectomy and study participation were achieved after full informed consents and written agreements between patients and clinicians in the surgical clinic. Entry criteria of the study included (1) both women and men, (2) patients aged 18-70 years, (3) with all patients (subjects) meeting the criteria for LSG following American Society for Metabolic and Bariatric Surgery guidelines^13^, a body mass index (BMI) of >40 kg/m^2^ or of >35 kg/m^2^ with at least one co-morbidity (e.g., type 2 diabetes (T2D), hypertension, or dyslipidemia). Patients with pre-existing autoimmune diseases or on immunomodulatory treatments were excluded.

Approval for the study was obtained from the Partners Human Research Committee (PHRC), the Institutional Review Board (IRB) of Partners HealthCare (IRB protocols 2015P000500 and 2015P000787).

### Study design

All subjects were recruited preoperatively within one month of surgery and followed up post-operatively at 3, 6, 9, and 12 months.

Prior to surgery, all subjects were required to complete a baseline questionnaire on clinical information and changes of parameters were monitored at subsequent time-points. Standard LSG comprised of a vertical stapled resection of the greater curvature of stomach over a 36-to 40-Fr bougie based on surgeon’s preference. Whole blood samples were collected peri-operatively (fasting) and at three (fasting), six, nine, and twelve months (fasting) following surgery. Patient demographics and clinical characteristics were recorded in an electronic database (REDCap, version 8.10.20)^14^ housed at Enterprise Research Information Science and Computing at Partners HealthCare.

Out of the 24 enrolled subjects, one participant dropped out of study prior to baseline (T0). 23 patients reached three months follow up (T3), 18 at six months (T6), 13 at nine months (T9) and 12 patients completed the full study at twelve months (T12).

### Measurement of clinical and biochemical data

At each timepoint, the following clinical information was ascertained by the means of administering a study questionnaire or consulting the electronic medical record; age, gender, race, past medical history, and BMI. All blood samples were collected on ice, centrifuged, and separated within two hours of collection and subsequently processed. Biochemical measurements were analyzed within the local hospital-accredited laboratory (Laboratory Corporation of America Holdings (Morrisville, NC) and Brigham Research Assay Core (Boston, MA)). A portion of the blood samples were stored in PAXgene Blood RNA Tubes (QIAGEN, Hilden, Germany) for later RNA-sequencing.

### Statistical analysis

Categorical data were presented as counts or percentages. Continuous variables with normal distribution were reported as means and bootstrapped non-parametric 95% confidence intervals with 1000 resamples (95% CI) or standard deviations (SD). *Post hoc* pair-wise comparisons of each follow-up time point versus baseline were performed using a paired Welch’s *t*-test. All tests were two-sided with the level of significance set as *P* < 0.05 and were corrected for multiple comparisons using the Holm method^15^. Statistical analyses were performed using GraphPad Prism (version 7.00) for Windows (GraphPad Software (La Jolla, CA)) or using R (version 3.6)^16^ and Bioconductor (version 3.10)^17,18^. Phenotype data were accessed programmatically using the R package “REDCapR” (version 0.9.8).

### Gene expression profiling

An RNA-sequencing (RNA-seq) pilot was performed on pairs of whole blood samples from the first six subjects of the study (baseline and three months post-op). Sequencing was performed at Partners HealthCare Personalized Medicine (Boston, MA). Three Illumina sequencing batches of 75 base-pair paired-end RNA-seq data in the FASTQ file format were generated across multiple lanes. The lanes were merged to create single-sample FASTQ files, which were trimmed with the Skewer software (version 0.1.118)^19^ and aligned to the GRCh38 genome reference assembly using the Spliced Transcripts Alignment to a Reference (STAR) software (version 2.5.2b)^20^. The aligned BAM files were sorted and quantified using the Python library “HTSeq” (version 0.6.1p1)^21^. The raw read counts were normalized using the R package “DESeq2” (version 1.26)^22^.

Differential gene expression (DGE) modeling was performed using DESeq2. In total, 65,986 distinct gene transcripts were assayed based on the reference transcriptome, but only 36,593 remained after filtering out transcripts with mean normalized read counts of zero across the 12 samples. Given the small sample size for this pilot analysis (*n* = 6), the paired-sample DGE model included only study timepoint as a predictor. Correction for multiple testing was made using Independent Hypothesis Weighting as implemented in the Bioconductor package “IHW” (version 1.14)^23,24^. DGE results were visualized using the R package “EnhancedVolcanoPlot” (version 1.4). Transcript counts were transformed by variance stabilization in DESeq2 and then bidirectional hierarchical clustering by Euclidean distance was performed on a subset of transcripts with significant DGE results using the R package “pheatmap” (version 1.0). Gene set enrichment analysis (GSEA) of the DGE results was performed using the Bioconductor package “fgsea” (version 1.12)^25^. Hallmark gene sets from the Molecular Signatures Database (version 6.0) were tested as GSEA hypotheses^26^. GSEA results were visualized using the Bioconductor package “clusterProfiler” (version 3.14)^27^.

## Results

### Baseline demographic characteristics

The baseline demographic data and preoperative co-morbidities are summarized in **Table 1**. The mean age (and SD) of the 23 subjects was 44.2 ± 12.3 years; 78% of the subjects were female; the subjects had a mean weight of 124.6 ± 21.1 kg and a BMI of 45.2 ± 7.2 kg/m^2^.

**Table 1.**
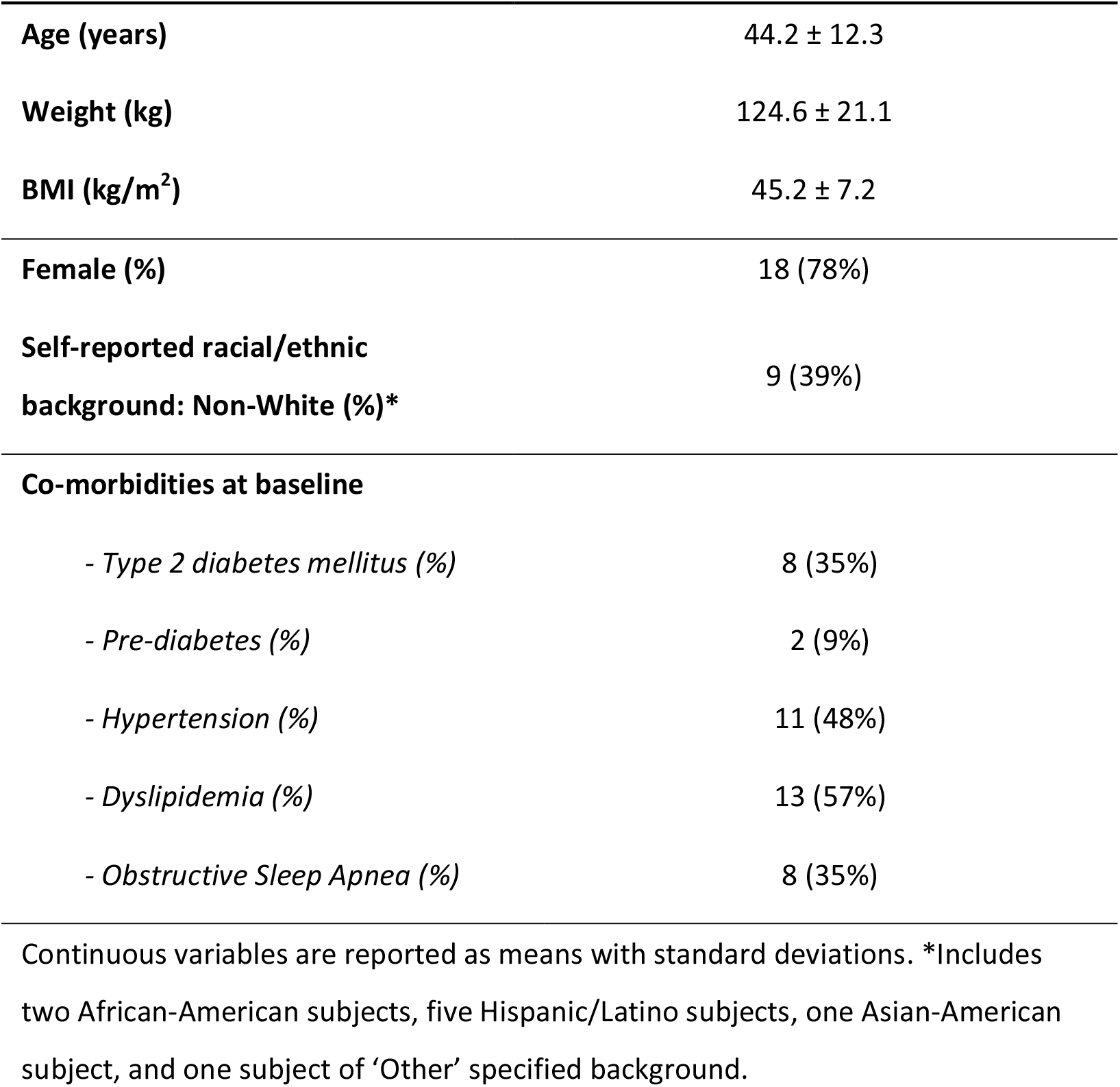
Pre-operative baseline demographics and obesity-related co-morbidities of 23 study subjects undergoing sleeve gastrectomy

|  |  |
| --- | --- |
| **Age (years)** | 44.2 ± 12.3 |
| **Weight (kg)** | 124.6 ± 21.1 |
| **BMI (kg/m<sup>2</sup>)** | 45.2 ± 7.2 |
| **Female (%)** | 18 (78%) |
| **Self-reported racial/ethnic background: Non-White (%)\*** | 9 (39%) |
| **Co-morbidities at baseline** |  |
| - *Type 2 diabetes mellitus (%)* | 8 (35%) |
| - *Pre-diabetes (%)* | 2 (9%) |
| - *Hypertension (%)* | 11 (48%) |
| - *Dyslipidemia (%)* | 13 (57%) |
| - *Obstructive Sleep Apnea (%)* | 8 (35%) |

Continuous variables are reported as means with standard deviations. \*Includes two African-American subjects, five Hispanic/Latino subjects, one Asian-American subject, and one subject of 'Other' specified background.

### Weight loss and related improvements in T2D

Weight and BMI were recorded every three months up to one year post-operatively. Significant reductions were observed in total body weight loss by three months post-op (T3) (mean = 18.1%, 95% CI: [16.6, 19.6], *P* < 0.001), six months (T6) (24.0%, 95% CI: [20.6, 27.2], *P* < 0.001), nine months (T9) (29.2%, 95% CI: [24.4, 34.3], *P* < 0.001), and twelve months (T12) (29.6%, 95% CI: [23.7, 35.4], *P* < 0.001) (**Figure 1A-B**). Loss to follow-up and/or missing data meant that sample size decreased by later time points (T6 to T12).

**Figure 1.**
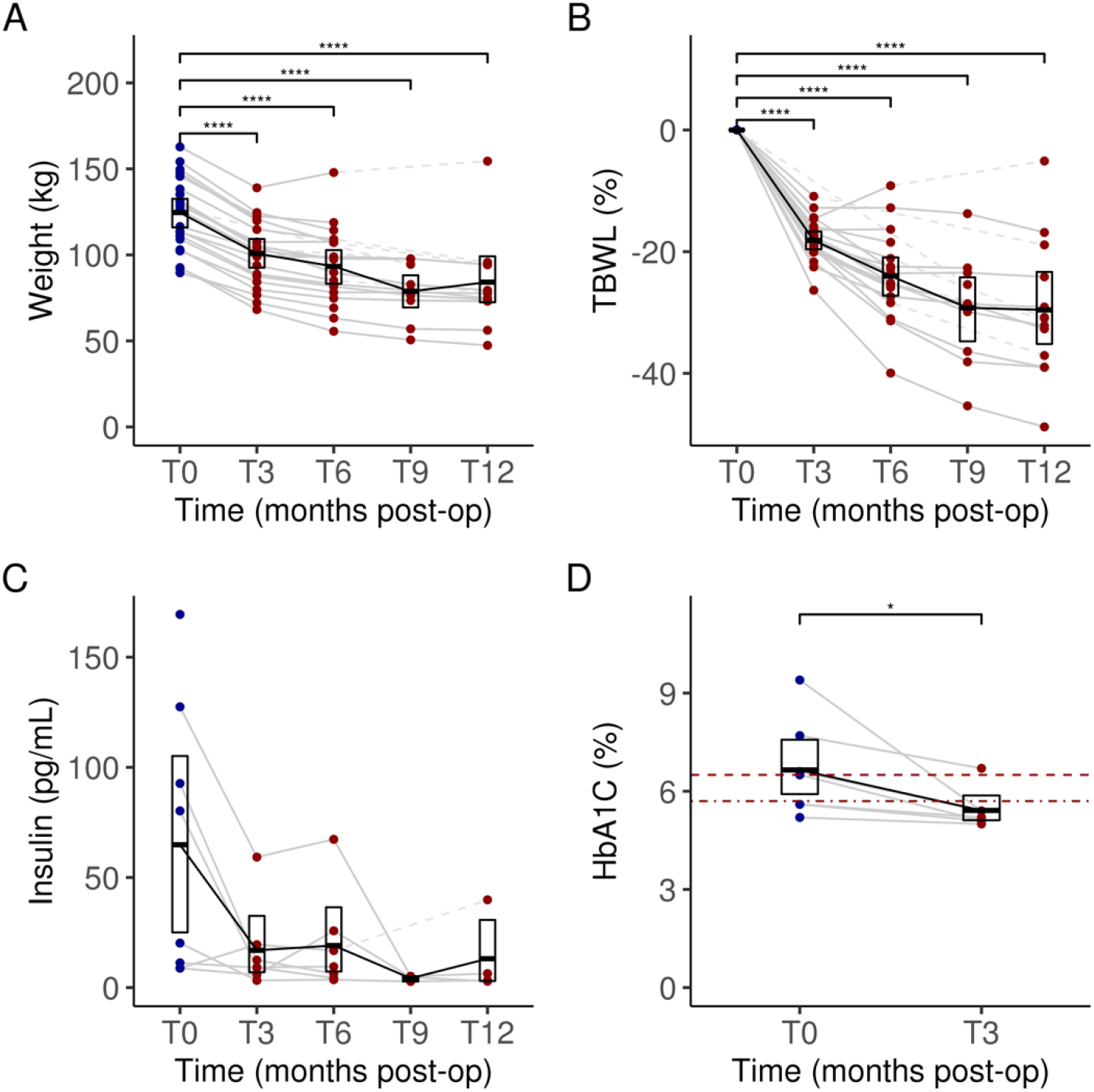
Weight change (**A**) and percent total body weight loss (TBWL) (**B**) for each subject over 12 months following sleeve gastrectomy. Serum insulin levels in subjects with T2D (**C**) over 12 months post operatively. Comparison of HbA1C (%) levels in subjects with T2D (**D**) at three months to baseline. Dark blue dots indicate values measured at baseline (pre-operatively) and dark red dots indicate values measured during follow-up (post-operatively). Dashed lines between dots indicate missing data for a given subject. Data for each timepoint are summarized as means and 95% confidence intervals. In panel **D**, the two dark red horizontal lines indicate the HbA1C thresholds for defining prediabetes (bottom, dot-dashed, HbA1C = 5.7%) and diabetes (top, dashed, HbA1C = 6.5%). Significant differences adjusted for multiple testing were marked with the following thresholds: ^*^*P* < 0.05; ^**^*P* < 0.01, ^***^*P* < 0.001, ^****^*P* < 0.0001. T0, baseline; T3, 3 months post-op; T6, 6 months; T9, 9 months; T12, 12 months.

In a subset of eight diabetic subjects, who had baseline preoperative mean hemoglobin A1c (HbA1c) values of at least 6.5% and/or were actively using T2D medications, we examined two relevant biomarkers. Among these diabetic subjects, mean fasting plasma insulin levels were trending lower following LSG, from 64.8 µIU/mL (95% CI: [28.5, 105.0]) at T0 to 13.1 µIU/mL (95% CI: [3.1, 30.7]) at T12 (**Figure 1C**), though this difference was not significant due to a large baseline variance (*P* ≥ 0.05). They also had a significant reduction and normalization of HbA1C by three months after LSG, changing from a pre-operative mean of 6.7% (95% CI: [5.9, 7.5]) to a post-operative mean of 5.4% (95% CI: [5.1, 5.9], *P* < 0.05) (**Figure 1D**).

### Leukocyte profiles and clinical markers of inflammation

As a sign of chronic inflammation, plasma clinical inflammatory markers were elevated at baseline prior to LSG, including white blood cells (WBC) (9.4 K/µL, 95% CI: [8.3, 10.6]), neutrophils (6.7 K/µL, 95% CI: [5.6, 7.7]), CRP (7.0 mg/L, 95% CI: [5.2, 9.1]) and IL-6 (5.4 pg/mL, 95% CI: [3.9, 7.2]). This obesity-related chronic inflammatory state was ameliorated throughout the 12 months with significant changes observed as early as T3. WBC levels were reduced to 6.7 K/µL (95% CI: [6.3, 7.2], *P* < 0.001) at T3 and 5.9 K/µL (95% CI: [5.2, 6.7], *P* < 0.05) at T12 (**Figure 2A**). Reductions were also observed at T3 for CRP (4.7 mg/L, 95% CI: [3.3, 6.1]) (**Figure 2B**) and IL-6 (4.3 pg/mL, 95% CI: [3.2, 5.7]) (**Figure 2C**), though the differences were not significant (both *P* ≥ 0.05). Further reductions occurred at T12 for CRP (2.4 mg/L, 95% CI: [1.2, 3.6], *P* < 0.05) and IL-6 (2.4 pg/mL, 95% CI: [1.7, 3.1], *P* ≥ 0.05).

**Figure 2.**
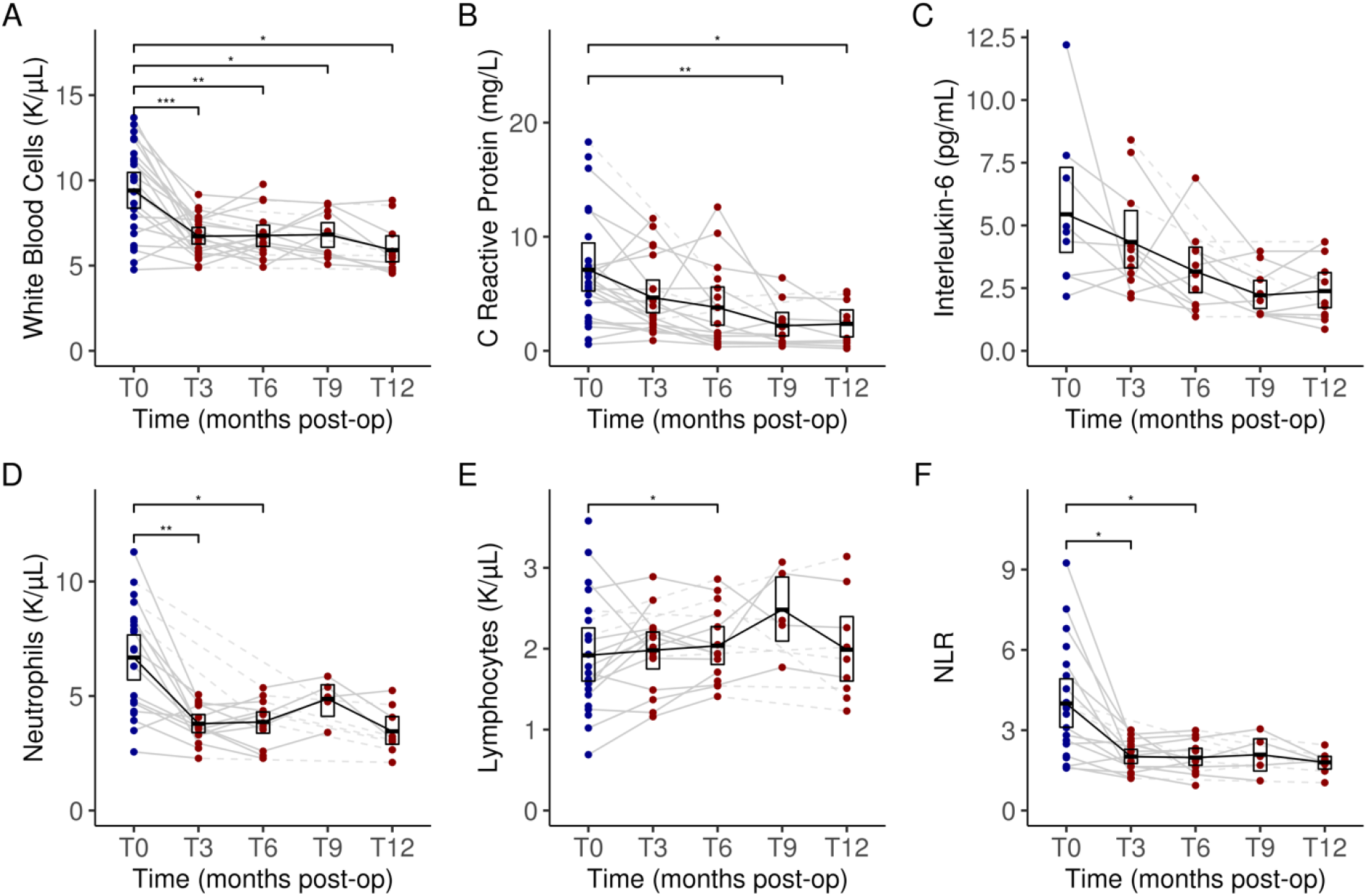
Changes in key serum clinical inflammatory markers and leukocytes composition: white blood cells (**A**), C reactive protein (**B**), interleukin-6 (**C**), neutrophil counts (**D**), lymphocyte counts (**E**) and neutrophil to lymphocyte ratio (**F**) from baseline to 12 months after undergoing sleeve gastrectomy. Dark blue dots indicate values measured at baseline (pre-operatively) and dark red dots indicate values measured during follow-up (post-operatively). Dashed lines between dots indicate missing data for a given subject. Data for each timepoint are summarized as means and 95% confidence intervals. Significant differences adjusted for multiple testing were marked with the following thresholds: ^*^*P* < 0.05; ^**^*P* < 0.01, ^***^*P* < 0.001, ^****^*P* < 0.0001. T0, baseline; T3, 3 months post-op; T6, 6 months; T9, 9 months; T12, 12 months.

Leukocyte composition was also altered post-LSG. A gradual reduction in neutrophil counts in plasma was observed from T0 (6.7 K/µL, 95% CI: [5.6, 7.7]) to T12 (3.5 K/µL, 95% CI: [2.9, 4.1]) (*P* ≥ 0.05) (**Figure 2D**) paired with a steady level of lymphocyte counts from T0 (1.9 K/µL, 95% CI: [1.6, 2.3]) to T12 (2.0 K/µL, 95% CI: [1.6, 2.5]) (*P* ≥ 0.05) (**Figure 2E**). Correspondingly, a reduction in neutrophil-lymphocyte ratio (NLR) was also observed from T0 (4.0, 95% CI: [3.1, 4.9]) to T12 (1.8, 95% CI: [1.6, 2.0]) (*P* ≥ 0.05) (**Figure 2F**). Importantly, the first significant reduction in NLR was observed as early as T3 (2.0, 95% CI: [1.7, 2.3], *P* < 0.05).

### Plasma hormonal biomarkers

Having observed a marked reduction in systemic inflammation following LSG, we evaluated the impact of the LSG on several biomarkers of adipose tissue-related inflammation (adiponectin and resistin) and hormonal biomarkers of metabolism (leptin and ghrelin). Although small improvements following LSG were observed for the measured hormonal plasma markers of adipose tissue-related inflammation, the changes were non-significant (all *P* > 0.05) and were not sustainable (**Figures 3A** and **3B**).

**Figure 3.**
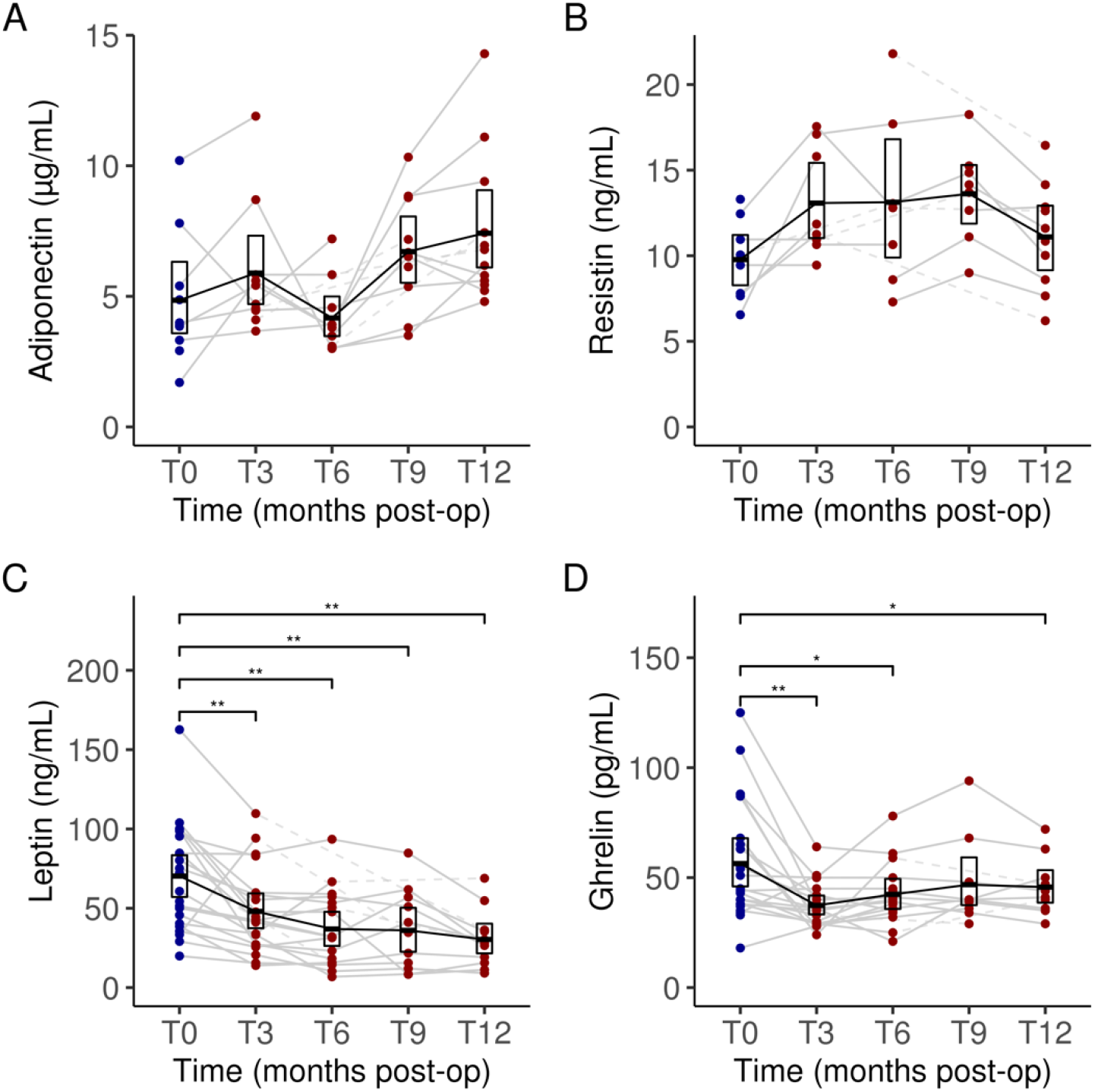
Changes in serum levels of adiponectin (**A**), resistin (**B**), leptin (**C**) and ghrelin (**D**) from baseline to 12 months after undergoing sleeve gastrectomy. Blue bars indicate values at baseline (preoperatively). Dark blue dots indicate values measured at baseline (pre-operatively) and dark red dots indicate values measured during follow-up (post-operatively). Dashed lines between dots indicate missing data for a given subject. Data for each timepoint are summarized as means and 95% confidence intervals. Significant differences adjusted for multiple testing were marked with the following thresholds: ^*^*P* < 0.05; ^**^*P* < 0.01, ^***^*P* < 0.001, ^****^*P* < 0.0001. T0, baseline; T3, 3 months post-op; T6, 6 months; T9, 9 months; T12, 12 months.

However, in contrast to the patterns observed for the tested inflammatory hormones, LSG was associated with marked and sustained improvements in the metabolic hormonal biomarkers (**Figures 3C** and **3D**). At three months (T3) following LSG, mean plasma concentrations of both leptin and ghrelin were reduced at all timepoints: leptin concentrations fell from 70.3 ng/mL (95% CI: [57.1, 84.4]) at baseline (T0) to 30.4 ng/mL (95% CI: [21.5, 39.8]) by T12 (*P* < 0.01); active ghrelin concentrations fell from 56.4 pg/mL (95% CI: [46.0, 67.9]) at T0 to 37.4 pg/mL (95% CI: [33.2, 42.4] at T3 (*P* < 0.01), 42.4 pg/mL (95% CI: [35.7, 49.8] at T6 (*P* < 0.05), and 45.7 pg/mL (95% CI: [38.4, 53.7]) at T12 (*P* < 0.05).

### Blood gene expression changes in several key metabolic pathways

To better understand how LSG improved immune and metabolic profiles, we evaluated whether any genome-wide changes in whole blood transcriptome profiling were associated with post-LSG weight loss. In the first six subjects from this cohort (four women and two men), we performed a pilot differential gene expression analysis to identify genes whose transcript counts were altered between baseline and three months following LSG. This subset of subjects had weight loss comparable to the rest of the cohort (TBWL = 19.4% vs. 17.6%). Of 36,593 gene transcripts detected, 2,557 (7.1%) demonstrated a significant change in expression three months after LSG compared to the baseline pre-operative state (false discovery rate (FDR) < 5%) (**Figure 4**), 337 of which showed more than 1 unit of log_2_ fold change (FC) (**Supplementary Table 1**).

**Figure 4.**
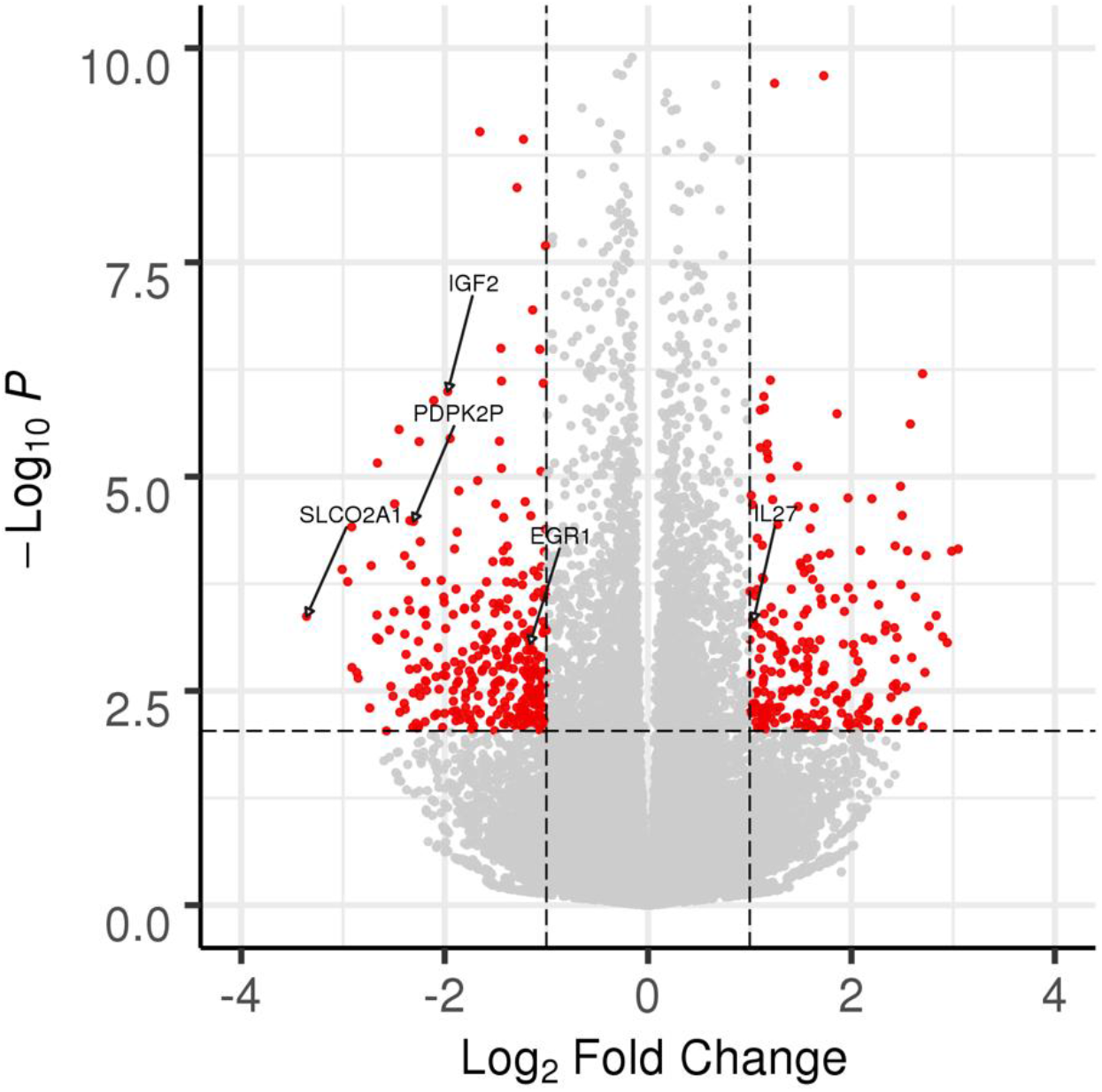
Differential expression of 36,593 gene transcripts in whole blood from six subjects at three months following sleeve gastrectomy. The dashed horizontal line corresponds to a false discovery rate (FDR) of 5% (corresponding to nominal *P* = 0.0093). The dashed vertical line corresponds to a log_2_ fold-change (FC) = 1.0. Transcripts showing significant and substantive differential expression (FDR < 5%, log_2_ FC > 1.0) are plotted in red. All other transcripts not meeting these two criteria are plotted in light gray. Transcripts of interest are annotated with their respective gene names and are described in the main text (*IL27, EGR1, SLCO2A1, IGF2, PDPK2P*).

We subsequently sought to explore whether LSG affects these transcriptomes in a similar manner by performing unsupervised hierarchical clustering on sample-specific transcript counts. We chose as input a set of 112 transcripts measured in these six individual subjects that showed strong enough differential expression to clearly differentiate pre-operative baseline (T0) samples from the 3-month post-operative (T3) samples (log_2_ FC > 1.635, FDR < 5%). While five of the six individual subjects clearly clustered together based on their baseline samples, we identified an outlier subject (LSG0006) whose baseline transcript expression levels were more similar to the set of follow-up samples (**Figure 5**). Interestingly, LSG0006 was also an outlier in weight loss response to LSG, achieving maximal weight loss at T3 (14.6% TBWL) and then experiencing early weight regain at T6 leading to the least weight loss in the cohort at T12 (5.1% TBWL) (**Figure 1B**).

**Figure 5.**
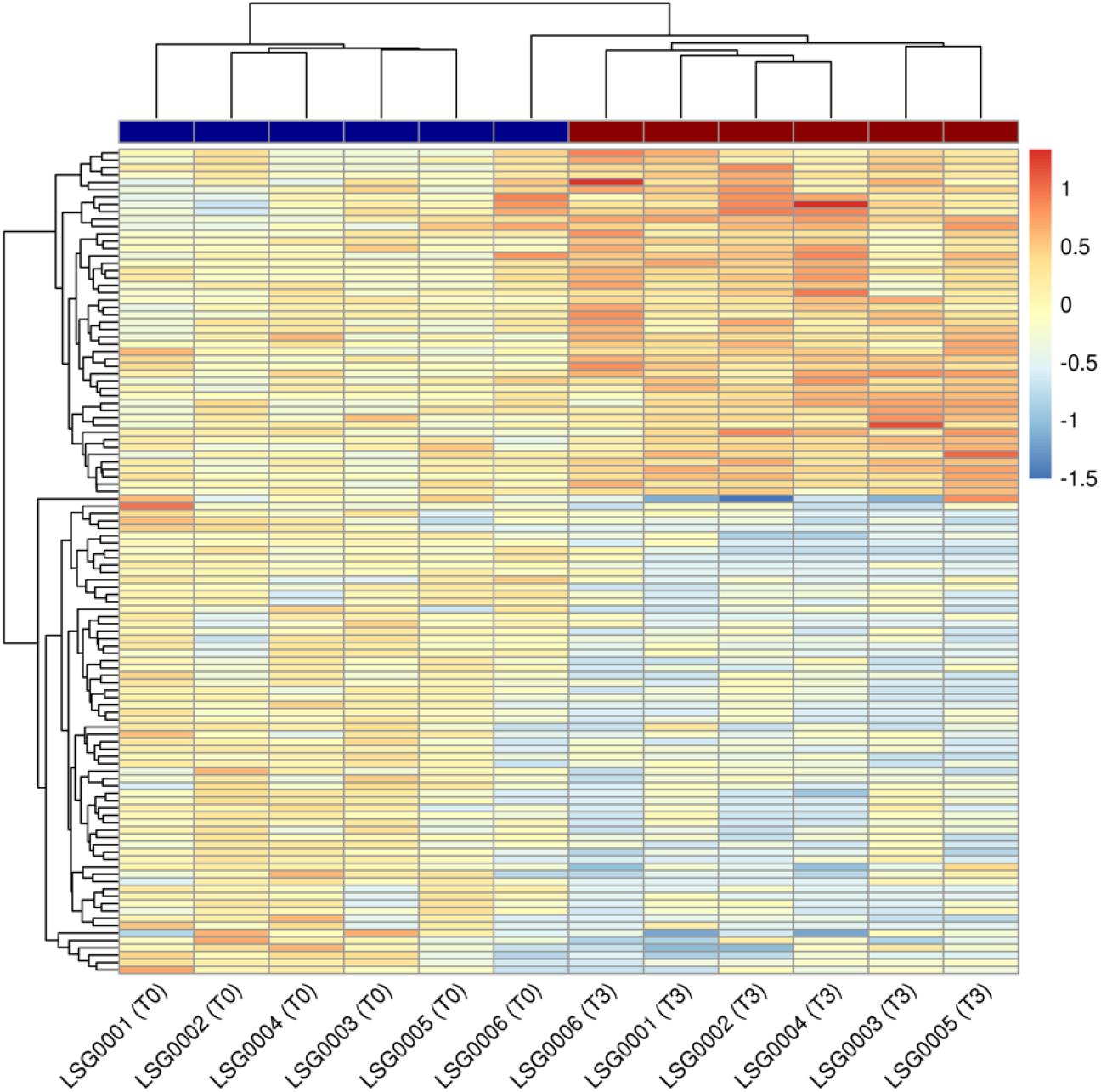
Heatmap of 112 individual transcript counts from strongest differential expression results in whole blood from six subjects at three months following sleeve gastrectomy (T3). Transcripts showing significant and substantive differential expression (false-discovery rate < 5%, log_2_ fold-change > 1.635) were chosen for display. All expression values are centered on the mean values across all six baseline (T0) samples. Red coloring in the heatmap indicates increased expression compared to the baseline mean (beige) across all subjects and blue indicates decreased expression. Samples (columns) are also annotated according to their timepoint: T0 (dark blue) and T3 (dark red).

Further exploration of the full subset of 405 significantly differentially expressed genes revealed several candidates of particular biological interest. First, we detected a change in the expression of genes involved in the inflammatory process following LSG, with a downregulation in pro-inflammatory mediators. We observed a reduction in expression of *EGR1*, implicated in the activation of T lymphocytes and macrophage differentiation^28^, as well as a downregulation of *SLCO2A1*, a cellular uptake transporter of prostaglandin E2, which stimulates the activation of proinflammatory cytokines^29,30^. In addition, LSG appears to induce an anti-inflammatory response, particularly with an upregulation of *IL27*, an anti-inflammatory cytokine involved in the production of interleukin-10^31,32^. Moreover, these immune cells appeared to undergo a series of metabolic and functional modifications. *IGF2*, a family of insulin-like growth factors, assumes major roles in growth and development associated with obesity^33^, was found to be downregulated following LSG. There was also a reduction in *PDPK2* gene expression, an important regulatory enzyme in glucose metabolism that controls pyruvate diversion to facilitate aerobic glycolysis^34^.

To assign broad biological meaning to these differential gene expression results, a set of 50 hallmark gene sets^26^, corresponding to distinct and coherent biological pathways, were tested for enrichment. In total, 28 gene sets were enriched at FDR < 5% (**Figure 6**). The gene sets represented a variety of biological pathways that again suggested a close relationship between immunity and cell metabolism in the context of surgical weight loss. Among the pathways most enriched for increased expression following surgery were related to oxidative phosphorylation, fatty acid oxidation, and Myc signaling; pathways that are essential to cell metabolic processes as well as cell proliferation, respectively.

**Figure 6.**
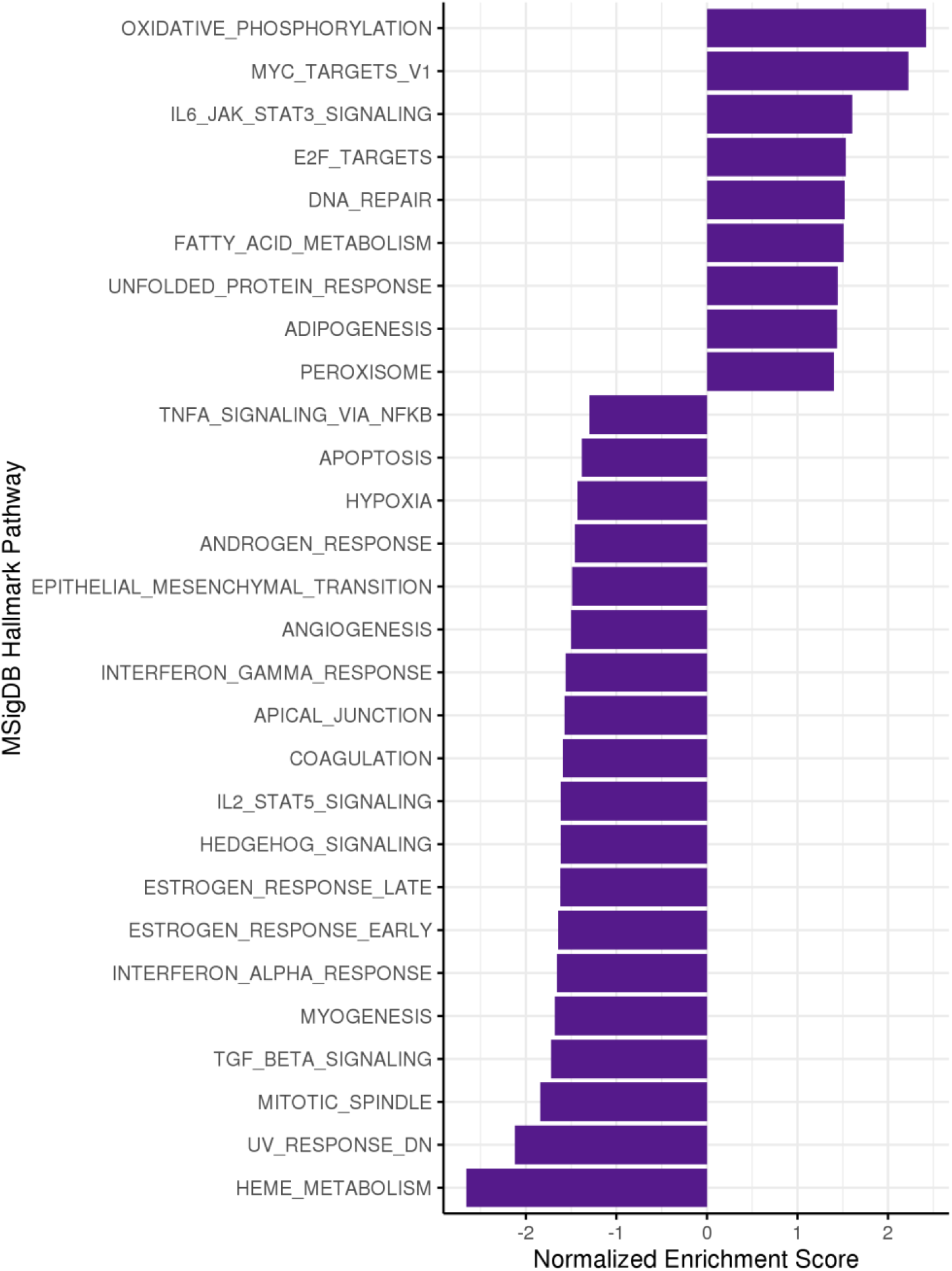
A set of 28 hallmark gene sets significantly enriched for differential expression in whole blood from six subjects at three months following sleeve gastrectomy at a false discovery rate (FDR) < 5%. Negative normalized enrichment scores indicate decreased expression of genes in the set following surgery, and positive score indicate increased expression. MSigDB, Molecular Signatures Database.

Concordantly, the gene sets most enriched for downregulated expression following surgery were also closely related to immune cell metabolic function. Pathways related to estrogen response and heme metabolism, both essential biological processes involved in signal transduction and immune cell survival were found to be downregulated post-LSG. Furthermore, a downregulation in pro-inflammatory pathways are once again reflected in our pathway analysis, with reduced gene expressions for transforming growth factor beta (TGFß) signaling and several cytokine pathways.

Collectively, our results demonstrated that the genes most enriched for both upregulation and downregulation represent metabolic pathways, suggesting that cellular metabolism plays an important role in supporting immune cell activation, maintenance, and differentiation.

## Discussion

The physiology and molecular underpinnings of the effects of laparoscopic sleeve gastrectomy on weight, metabolism, and immune function are likely far more complex than anatomical modifications alone. In this prospective, longitudinal study, we (1) characterized the temporal changes in serum biomarkers over a year; (2) characterized concurrent temporal changes in white blood cell composition; and (3) identified near-term gene expression alterations in several key immune and metabolic pathways in whole blood following LSG.

We demonstrated profound changes in hormonal markers including insulin, HbA1C, ghrelin, and leptin. Rapid improvements in fasting insulin concentrations and signs of diabetes remission has been reported within days following LSG^35^. Mounting evidence suggests that the significant improvement in insulin resistance is a result of a combination of weight-independent effect secondary to metabolic hormones as well as subsequent weight loss. Gut hormones, such as ghrelin and leptin, play an integral role in the appetite-signaling process have also been shown to significantly decrease following LSG^36^.

Additionally, we observed the recovery of the immune system from its chronic inflammatory state. Our findings are in concordance with previous studies demonstrating improvements in metabolic hormones and circulating inflammatory cytokines following bariatric surgery^11,37,38^ and weight loss^39^. Nonetheless, our present work offers a unique perspective by sampling patients at five evenly spaced perioperative time points, which gave us a more complete appreciation of the temporal changes associated with LSG. Intriguingly, most of the improvements in clinical metabolic and immune function biomarkers dramatically took place by three months following LSG, with sustainable results at one year. We acknowledge that these modifications may have even started earlier post operatively as suggested in several studies^35,38^.

The composition of leukocytes was also altered following surgery, with a reduction in absolute neutrophil counts. The differentiation of these leukocyte subpopulations may vary based on their role in the inflammatory response, as neutrophil activation is aligned with an active innate immune system, while lymphocytes are associated with the adaptive immune system^40^. The intricate rebalance of neutrophil and lymphocytes following LSG amount to a reduced NLR, signifying a decrease in the detrimental effects of neutrophilia and a transition to a more lymphocyte-mediated adaptive immunity. Along with other clinical inflammatory markers, there is an emerging interest in NLR for its prognostic value. A high postoperative day-one NLR is shown to a predictor for 30-day complications including longer hospital stay, major complications, and re-operation rates^41^.

We then performed RNA sequencing on human whole blood prior to and three months after LSG, to comprehensively characterize gene expression changes that may underlie LSG effects. Pathway analysis revealed robust changes in gene expression influencing both immune function and immune metabolism in nearly all subjects. At baseline, gene expression patterns in the subjects were also overall quite similar, with the exception of a single subject. This outlier, LSG0006, was a poor responder to LSG, having experienced both weight regain and failure to achieve diabetes remission. Interestingly, their baseline gene expression profile was more closely related to the post-LSG expression profile of the other subjects. The power of whole blood gene expression profiles for predicting response to LSG merits further study.

In terms of possible biological mechanisms, we found evidence of substantial upregulations of oxidative phosphorylation (OXPHOS) and fatty acid oxidation (FAO) metabolic pathways. Cellular metabolism has been suggested to play a critical role in immune cell reprogramming affecting macrophage, B and T cell activation, and proliferation^42–44^. In particular, a shift towards OXPHOS and FAO is linked to a regulatory, anti-inflammatory phenotype in multiple cellular immune subsets^45^. For example, pro-inflammatory M1 macrophages rely mainly on aerobic glycolysis to meet their ATP requirements. In contrast, M2 cells, which offer an anti-inflammatory profile, are more dependent on OXPHOS and FAO to provide substrates for the complexes of the electron transport chain^42,43,46^. As pro-inflammatory cells including M1 macrophages directly contribute to insulin resistance, an immunometabolic shift to oxidative phosphorylation and fatty acid oxidation is a potential mechanism by which LSG improves immune function and insulin resistance.

In addition, there is a complex interplay between immune markers following surgery. We observed a general downregulation in cytokine pathways along with an upregulation in gene expressions of several immunoglobulins. This is in line with a number of human and animals studies that have also demonstrated a rise in circulating and adipose tissue immunoglobulins along with a change the regulatory function of B lymphocytes following bariatric surgery^47,48^. These modifications in the adaptive immune system may explain the improvements in autoimmune or immune-mediated diseases detected following bariatric surgery^35,49^.

Lastly, we have also identified several metabolic-centric pathways, such as estrogen and heme metabolism, that are downregulated post-LSG. Estrogen receptors play a pivotal role in the regulation of innate immune signaling pathways and myeloid cell development^50^. Overexpression of heme metabolism may contribute to reactive oxygen species production, cellular hypoxia and DNA damage leading to cell death, a phenomenon often observed in chronic inflammation, obesity, and T2D^51,52^.

Taken together, our data suggest that LSG may have a profound effect on immune cells through the modulation of their own metabolic processes. We have previously published data that describes changes within metabolic and immune pathways in visceral adipose tissue following LSG using rodent model^47^. However, to our knowledge, this complex post-LSG relationship between immunity remodeling and cell metabolism within human immune cells has not been previously described.

Our study has several limitations. While the study was prospective in nature, we only analyzed subjects who elected to undergo a single type of bariatric surgery procedure. The sex distribution of the cohort was skewed towards female, characteristic of a bariatric surgery cohort^8^. We have a smaller data set for longer time points; however, this was not due to poor follow-up rate but rather reflects the ongoing development of a prospective study cohort. Despite having a satisfactory follow-up rate, several data points are missing due to hemolyzed blood samples or inadequate blood sampling. Our pilot transcriptomic study was also relatively modest in size, comprising six subjects, and was performed using whole blood, which is both a complex mélange of cell types and an indirect readout of metabolic programming throughout the body. Nonetheless, our data suggest several noteworthy pathways meriting future investigations in obese human subjects undergoing bariatric surgery.

In summary, we find sleeve gastrectomy induces marked early changes in immune cell composition, function, and metabolism, as measured by biomarkers and gene expression. Further evaluation is required of bariatric surgery’s effects on immunometabolism and consequences for vaccine response, host defense against pathogens, and allergy development. These studies may benefit from measurement of circulating immune cell function and health, together with their impact on metabolism. Identifying and understanding these associations will allow for the implementation of specific therapeutic strategies in obesity and its metabolic comorbidities.

## Data Availability

The phenotype and gene expression data described in this study can only be shared with IRB-approved investigators within Partners HealthCare.

## Author Contributions

- A.T. and E.G.S. performed the surgeries.
- D.C.C-C., B.A.R., A.T., and E.G.S. designed this work.
- D.C.C-C., B.A.R., and E.G.S. funded this work.
- D.C.C-C. and E.G.S. co-supervised this work.
- D.C.C-C., E.M.L., A.M.W., O.J.I.-B., C.O.I., E.J.B., and E.G.S. performed the study recruitment and clinical phenotyping.
- T.L., E.J.M.R., R.S., K.H., S.T.M., L.S., and L.F. processed the samples and/or assayed the biomarkers.
- T.L. and D.C.C-C. performed the statistical analyses.
- R.P.C. and D.C.C-C. developed and ran the gene expression analysis pipeline.
- T.L., D.C.C-C., and E.G.S. drafted the manuscript.
- All co-authors read and approved this work.

## Acknowledgements

The authors thank the study subjects for their generous participation in this work.

## References

1. Xu H, Barnes GT, Yang Q, et al. Chronic inflammation in fat plays a crucial role in the development of obesity-related insulin resistance. J Clin Invest. 2003;112(12):1821–1830. doi:10.1172/JCI19451

2. Hotamisligil GS, Shargill NS, Spiegelman BM. Adipose expression of tumor necrosis factor- (alpha): Direct. Science (80-). 1993;259(5091):87–92.

3. Hotamisligil GS. Inflammation, metaflammation and immunometabolic disorders. Nature. 2017;542(7640):177–185. doi:10.1038/nature21363

4. Weisberg SP, Leibel RL, Anthony W, et al. Obesity is associated with macrophage accumulation in adipose tissue Find the latest version : Obesity is associated with. J Clin Invest. 2003;112(12):1796–1808. doi:10.1172/JCI200319246.Introduction

5. Tse L. Chronic Inflammation in Fat Plays a Crucial Role in Development of Obesity. Screen. 2003;112(12):1821–1830. doi:10.1172/JCI200319451.Introduction

6. Lemieux I, Pascot A, Prud D, et al. Atherosclerosis and Lipoproteins Elevated C-Reactive Protein Another Component of the Atherothrombotic Profile of Abdominal Obesity. Atheroscler Lipoproteins. 2001;21:961–967.

7. GLOBAL STATUS REPORT on Noncommunicable Diseases 201 4 “Attaining the Nine Global Noncommunicable Diseases Targets; a Shared Responsibility.”

8. Schauer PR, Bhatt DL, Kirwan JP, et al. Bariatric surgery versus intensive medical therapy for diabetes - 5-year outcomes. N Engl J Med. 2017;376(7):641–651. doi:10.1056/NEJMoa1600869

9. Peterli R, Wölnerhanssen BK, Peters T, et al. Effect of Laparoscopic Sleeve Gastrectomy vs Laparoscopic Roux-en-Y Gastric Bypass on Weight Loss in Patients With Morbid Obesity: The SM- BOSS Randomized Clinical Trial. Jama. 2018;319(3):255–265. doi:10.1001/jama.2017.20897

10. Subramaniam R, Aliakbarian H, Bhutta HY, Harris DA, Tavakkoli A, Sheu EG. Sleeve Gastrectomy and Roux-en-Y Gastric Bypass Attenuate Pro-inflammatory Small Intestinal Cytokine Signatures. Obes Surg. 2019. doi:10.1007/s11695-019-04059-0

11. Mallipedhi A, Prior SL, Barry JD, Caplin S, Baxter JN, Stephens JW. Changes in inflammatory markers after sleeve gastrectomy in patients with impaired glucose homeostasis and type 2 diabetes. Surg Obes Relat Dis. 2014;10(6):1123–1128. doi:10.1016/j.soard.2014.04.019

12. Hakeam HA, O’Regan PJ, Salem AM, Bamehriz FY, Eldali AM. Impact of laparoscopic sleeve gastrectomy on iron indices: 1 Year follow-up. Obes Surg. 2009;19(11):1491–1496. doi:10.1007/s11695-009-9919-2

13. Mechanick J, Youdim A, Jones D, et al. Clinical Practice Guidelines for the Perioperative Nutritional, Metabolic, and Nonsurgical Support of the Bariatric Surgery Patient—2013 Update: Cosponsored by American Association of Clinical Endocrinologists, The Obesity Society, and American Society for Metabolic & Bariatric Surgery. 2013. doi:10.1016/j.soard.2012.12.010

14. Harris PA, Taylor R, Thielke R, Payne J, Gonzalez N, Conde JG. Research electronic data capture (REDCap)-A metadata-driven methodology and workflow process for providing translational research informatics support. J Biomed Inform. 2009;42(2):377–381. doi:10.1016/j.jbi.2008.08.010

15. Holm S. A Simple Sequentially Rejective Multiple Test Procedure. Scand J Stat. 2010;6(2):65–70. doi:10.2307/4615733

16. Hornik K. The R FAQ. The R FAQ. doi:10.1071/HEv24n3toc

17. Gentleman RC, Carey VJ, Bates DM, et al. Open Access Bioconductor: Open Software Development for Computational Biology and Bioinformatics. Vol 5.; 2004. http://genomebiology.com/2004/5/10/ http://genomebiology.com/2004/5/10/R80. Accessed October 10, 2019.

18. Huber W, Carey VJ, Gentleman R, et al. Orchestrating high-throughput genomic analysis with Bioconductor HHS Public Access. Nat Methods. 2015;12(2):115–121. doi:10.1038/nmeth.3252

19. Jiang H, Lei R, Ding S-W, Zhu S. Skewer: A Fast and Accurate Adapter Trimmer for next-Generation Sequencing Paired-End Reads. Vol 15.; 2014. http://www.biomedcentral.com/1471-2105/15/182. Accessed October 10, 2019.

20. Dobin A, Davis CA, Schlesinger F, et al. Sequence analysis STAR: ultrafast universal RNA-seq aligner. 2013;29(1):15–21. doi:10.1093/bioinformatics/bts635

21. Anders S, Pyl PT, Huber W. Genome analysis HTSeq-a Python framework to work with high-throughput sequencing data. 2015;31(2):166–169. doi:10.1093/bioinformatics/btu638

22. Love MI, Huber W, Anders S. Targeted analysis of nucleotide and copy number variation by exon capture in allotetraploid wheat genome. Genome Biol. 2011;15:550. doi:10.1186/s13059-014-0550-8

23. Ignatiadis N, Huber W. Covariate powered cross-weighted multiple testing. January 2017. http://arxiv.org/abs/1701.05179. Accessed May 30, 2020.

24. Ignatiadis N, Klaus B, Zaugg JB, Huber W. Data-driven hypothesis weighting increases detection power in genome-scale multiple testing. Nat Methods. 2016;13(7):577–580. doi:10.1038/nmeth.3885

25. Sergushichev AA. An algorithm for fast preranked gene set enrichment analysis using cumulative statistic calculation. doi:10.1101/060012

26. Liberzon A, Birger C, Thorvaldsdóttir H, Ghandi M, Mesirov JP, Tamayo P. The Molecular Signatures Database (MSigDB) hallmark gene set collection HHS Public Access. Cell Syst. 2015;1(6):417–425. doi:10.1016/j.cels.2015.12.004

27. Yu G, Wang LG, Han Y, He QY. ClusterProfiler: An R package for comparing biological themes among gene clusters. Omi A J Integr Biol. 2012;16(5):284–287. doi:10.1089/omi.2011.0118

28. Seiler MP, Mathew R, Liszewski MK, et al. Elevated and sustained expression of the transcription factors Egr1 and Egr2 controls NKT lineage differentiation in response to TCR signaling. Nat Immunol. 2012;13(3):264–271. doi:10.1038/ni.2230

29. Kanai N, Lu R, Satriano JA, Bao Y, Wolkoff AW, Schuster VL. Identification and Characterization of a Prostaglandin Transporter. Science (80-). 1995;268(5212):866–869. doi:10.2307/2887969

30. Shimada H, Nakamura Y, Nakanishi T, Tamai I. OATP2A1/SLCO2A1-mediated prostaglandin E2 loading into intracellular acidic compartments of macrophages contributes to exocytotic secretion. Biochem Pharmacol. 2015;98(4):629–638. doi:10.1016/j.bcp.2015.10.009

31. Murugaiyan G, Mittal A, Lopez-Diego R, Maier LM, Anderson DE, Weiner HL. IL-27 Is a Key Regulator of IL-10 and IL-17 Production by Human CD4 + T Cells. J Immunol. 2009;183(4):2435–2443. doi:10.4049/jimmunol.0900568

32. Ip WKE, Hoshi N, Shouval DS, Snapper S, Medzhitov R. Anti-inflammatory effect of IL-10 mediated by metabolic reprogramming of macrophages. Science (80-). 2017;356(6337):513–519. doi:10.1126/science.aal3535

33. Haywood NJ, Slater TA, Matthews CJ, Wheatcroft SB. The insulin like growth factor and binding protein family: Novel therapeutic targets in obesity & diabetes. Mol Metab. 2019;19:86–96. doi:10.1016/j.molmet.2018.10.008

34. Tan Z, Xie N, Cui H, et al. Pyruvate Dehydrogenase Kinase 1 Participates in Macrophage Polarization via Regulating Glucose Metabolism. J Immunol. 2015;194(12):6082–6089. doi:10.4049/jimmunol.1402469

35. Heshmati K, Lo T, Tavakkoli A, Sheu E. Short-Term Outcomes of Inflammatory Bowel Disease after Roux-en-Y Gastric Bypass vs Sleeve Gastrectomy. J Am Coll Surg. 2019;228(6):893-901.e1. doi:10.1016/j.jamcollsurg.2019.01.021

36. Dimitriadis E, Daskalakis M, Kampa M, Peppe A, Papadakis JA, Melissas J. Alterations in Gut Hormones After Laparoscopic Sleeve Gastrectomy. Ann Surg. 2013;257(4):647–654. doi:10.1097/SLA.0b013e31826e1846

37. Illán-Gómez F, Gonzálvez-Ortega M, Orea-Soler I, et al. Obesity and inflammation: Change in adiponectin, C-reactive protein, tumour necrosis factor-alpha and interleukin-6 after bariatric surgery. Obes Surg. 2012;22(6):950–955. doi:10.1007/s11695-012-0643-y

38. Miller GD, Nicklas BJ, Fernandez A. Serial changes in inflammatory biomarkers after Roux-en-Y gastric bypass surgery. Surg Obes Relat Dis. 2011;7(5):618–624. doi:10.1016/j.soard.2011.03.006

39. Manigrasso MR, Ferroni P, Santilli F, et al. Association between circulating adiponectin and interleukin-10 levels in android obesity: Effects of weight loss. J Clin Endocrinol Metab. 2005;90(10):5876–5879. doi:10.1210/jc.2005-0281

40. Howard R, Kanetsky PA, Egan KM. Exploring the prognostic value of the neutrophil-to-lymphocyte ratio in cancer. Sci Rep. 2019;9(1):1–10. doi:10.1038/s41598-019-56218-z

41. Da Silva M, Cleghorn MC, Elnahas A, Jackson TD, Okrainec A, Quereshy FA. Postoperative day one neutrophil-to-lymphocyte ratio as a predictor of 30-day outcomes in bariatric surgery patients. Surg Endosc. 31. doi:10.1007/s00464-016-5278-y

42. Viola A, Munari F, Sánchez-Rodríguez R, Scolaro T, Castegna A. The metabolic signature of macrophage responses. Front Immunol. 2019;10(JULY). doi:10.3389/fimmu.2019.01462

43. Jung J, Zeng H, Horng T. Metabolism as a guiding force for immunity. Nat Cell Biol. 2019;21(1):85–93. doi:10.1038/s41556-018-0217-x

44. Jellusova J. Metabolic control of B cell immune responses. Curr Opin Immunol. 2020;63:21–28. doi:10.1016/j.coi.2019.11.002

45. Olenchock BA, Rathmell JC, Vander Heiden MG. Biochemical Underpinnings of Immune Cell Metabolic Phenotypes. Immunity. 2017;46(5):703–713. doi:10.1016/j.immuni.2017.04.013

46. Lehrke M, Lazar MA. Inflamed about obesity. Nat Med. 2004;10(2):126–127. doi:10.1038/nm0204-126

47. Harris DA, Mina A, Cabarkapa D, et al. Sleeve Gastrectomy enhances glucose utilization and remodels adipose tissue independent of weight loss. Am J Physiol Metab. February 2020. doi:10.1152/ajpendo.00441.2019

48. Dai X, Zhao W, Zhan J, et al. B cells present skewed profile and lose the function of supporting T cell inflammation after Roux-en-Y gastric bypass. Int Immunopharmacol. 2017;43:16–22. doi:10.1016/j.intimp.2016.11.033

49. Rudge EJM, Sheu EG. Bariatric surgery for autoimmunity? JAMA Surg. 2017;152(4):349–350. doi:10.1001/jamasurg.2016.4645

50. Kovats S. Estrogen receptors regulate innate immune cells and signaling pathways. Cell Immunol. 2015;294(2):63–69. doi:10.1016/j.cellimm.2015.01.018

51. Vijayan V, Wagener FADTG, Immenschuh S. The macrophage heme-heme oxygenase-1 system and its role in inflammation. Biochem Pharmacol. 2018;153:159–167. doi:10.1016/j.bcp.2018.02.010

52. Moreno-Navarrete JM, Rodríguez A, Ortega F, et al. Increased adipose tissue heme levels and exportation are associated with altered systemic glucose metabolism. Sci Rep. 2017;7(1):1–9. doi:10.1038/s41598-017-05597-2

